# “That is why I trust”: A qualitative study on acceptability and feasibility of novel tongue swab diagnostics to assess people presenting with tuberculosis symptoms in Viet Nam and Zambia

**DOI:** 10.1101/2025.06.30.25330535

**Authors:** Alyssa Sales, Hien Le, Seke Muzazu, Evelyn Kunda-Ng’andu, Maria del Mar Castro, Andrew D. Kerkhoff, Ha Phan, Claudia M. Denkinger, Adithya Cattamanchi, Nora West, Monde Muyoyeta

## Abstract

Millions of tuberculosis (TB) cases are estimated to be undiagnosed and unreported annually. Sputum has been the primary approach for diagnostic testing but tongue swabs are being investigated as an alternative to expand testing. To understand potential uptake and implementation, we explored the acceptability, usability, and feasibility of tongue swab-based TB testing from the perspective of healthcare workers, people undergoing TB screening, and caregivers in Viet Nam and Zambia.

We interviewed people with symptoms of TB, caregivers of children undergoing TB evaluation, and healthcare workers who collected tongue swabs (n = 76 participants) between September 2023 and February 2024. Interviews were analyzed using framework analysis to elucidate preferences, experiences, and acceptability of tongue swabs vs. sputum. Findings were further organized according to acceptability and feasibility to understand barriers and facilitators to uptake.

Most participants preferred tongue swab to sputum collection. The perceived usability and feasibility of tongue swabs were high. Key themes that influenced the acceptability of tongue swabs included ease of use, diagnostic accuracy, diagnostic yield, hygiene, risk of TB transmission during sample collection, time to test result, and trust in healthcare workers and the health system. Across interviews, many participants described tongue swabs as a comfortable and easy way to test for TB, compared to the physical discomfort and difficulty expectorating sputum. Participants described tongue swab suitability for everyone, yet perceived diagnostic accuracy was crucial in shaping test preference.

Tongue swab-based testing for TB is likely to be highly acceptable and feasible if incorporated into TB diagnostic guidelines.

## 1. Introduction

Tuberculosis (TB) persists as a leading cause of mortality globally, disproportionately affecting low- and middle-income countries (Tuberculosis, 2024). In 2023, 10.8 million people were diagnosed with TB, and approximately 1.25 million people died from TB. Many people with TB are not identified, with estimates indicating a diagnostic gap of 2.6 million people (Global Tuberculosis Report 2024).

Delayed and missed diagnoses remain a critical challenge along the TB care cascade. Key populations like children or those with early TB illness may experience difficulty producing sputum (Codsi et al., 2023) and providing a quality sample (Sakundarno et al., 2009). Sputum samples can vary in quality (Regati et al., 2021), impacting test accuracy. Additionally, sputum expectoration may pose safety hazards to healthcare workers and others seeking TB care (Andama et al., 2022; Codsi et al., 2023). Understanding the potential of alternative sample types to expand testing and lead to early TB detection and treatment is critical (High Priority Target Product Profiles for New Tuberculosis Diagnostics: Report of a Consensus Meeting, 2014; Target Product Profile for Tuberculosis Diagnosis and Detection of Drug Resistance, 2024).

Tongue swabs have emerged as a promising option for TB diagnostic testing. Tongue swabs may be easier to collect and enable faster results than sputum-based TB testing (Wood et al., 2024). Although previous studies have evaluated the sensitivity and specificity of tongue swabs (Andama et al., 2022; Church et al., 2024), studies exploring the perspectives related to tongue-swab-based TB testing, including acceptability and preferences of affected individuals, are lacking. Codsi et al. (2023) assessed healthcare workers’ attitudes towards tongue swabs, yet no other qualitative studies have evaluated tongue swabs as a TB diagnostic specimen. Studies are therefore needed to determine whether this novel specimen is likely to be trusted and accepted by people undergoing TB testing, their caregivers and healthcare workers.

To address this, we sought to understand the acceptability and feasibility of tongue swab-based TB testing and stakeholder preferences for use compared to sputum-based TB testing. Using qualitative methods, we explored the perceptions of people accessing TB evaluation, caregivers of children undergoing TB evaluation, and healthcare workers involved in TB care in Viet Nam and Zambia.

## 2. Methods

### 2.1 Study Setting and Population

From September 2023-February 2024, we conducted semi-structured in-depth interviews (IDIs) with adults (age 18 and above), caregivers of children (less than 13 years of age) undergoing TB evaluation, and healthcare workers who collected tongue swabs, at six primary health centers in Viet Nam and Zambia (Appendix A). Participants were recruited from primary health centers as they sought care. Our study was nested within a parent study (Tongue Swab Tuberculosis Diagnostic Yield [TSwaY]) evaluating the diagnostic yield of tongue swab-vs. sputum-based molecular testing for TB. Within the parent TSwaY study, participants with presumptive TB (based on symptoms or a positive TB screening test) were approached for recruitment and collection of tongue swab and sputum samples. We purposively sampled people with presumptive TB and caregivers of children with presumptive TB to achieve balance in age, gender, and, where applicable, refusal or acceptance of sputum and/or tongue swab collection. Sampling on these characteristics was chosen to explore similarities and differences in specimen collection experiences across relevant groups. All healthcare workers who engaged in tongue swab and sputum collection as part of the TSwaY study were also invited to participate.

### 2.2 Procedures

IDIs were conducted by experienced female Zambian and Vietnamese qualitative data collectors who received additional refresher qualitative interviewing training and practicing specific to the study objectives from the research team in advance of recruitment and data collection. All participants or their caregivers were provided written informed consent. Interviews were conducted in person at the health facilities following specimen provision for TB testing. At the time of the interview, participants did not yet have their TB test results. All IDIs were completed at a single time point and lasted approximately 30-60 minutes.

Semi-structured interview guides (Appendix B) were used to guide IDIs. Participants were asked about their experiences at the clinic for TB evaluation and their thoughts on tongue swab and sputum collection. IDI guides were developed based on research objectives and the theoretical framework of health care interventions (Martin et al., 2022; Sekhon et al., 2017). ID Is explored participant perceptions, the usability, defined as the ease of collecting and receiving a tongue swab sample, and the acceptability of these two specimen types for TB testing. All participants were asked about their likes and dislikes for each specimen type, if applicable, preference for sputum vs. tongue swab collection, and factors that influence their perception of tongue swab-based TB testing.

Individuals undergoing TB screening were additionally asked about their physical experiences during specimen collection and their thoughts on the instructions provided. From the consultation on TB evaluation with healthcare workers, and direct support and observation of their child’s experiences in the sample collection process, caregivers were asked to share their considerations for the acceptability of tongue swab (including their child’s perceived physical experience), compared to sputum-based tests, following the specimen provision. Healthcare workers were also asked about their perceptions of the preferences of individuals seeking care and how replacing sputum collection with tongue swab collection would impact usability and their daily work. Interview guides were developed in English and translated to Bemba and Nyanja for Zambian participants and Vietnamese for Vietnamese participants by bilingual research team members through an initial translation and back translation process, followed by team-based review and discussion before arriving at the final versions of the interview guides. All interviews were audio recorded, transcribed verbatim, and translated into English. The principle of saturation, when new or additional data does not yield new information, was used to arrive at the final sample size among people presenting for presumptive TB and caregivers (Sandelowski, 1995), with further targeted sampling conducted in Zambia to account for heterogeneity in acceptance, refusal, or inability to complete sputum or tongue swab collection. The sample size for healthcare workers included the number of healthcare workers across the study sites who engaged in tongue swab sample collection. All who participated in sample collection consented to participation. During data collection, study investigators met weekly with the data collection team to discuss interview progress and content and review debrief forms, notes, and available interview transcripts to assess saturation throughout data collection.

### 2.3 Analysis

A modified framework analysis was used to analyze the resulting qualitative data (Gale et al., 2013). Based on the interview guide, the study analysis team developed an initial codebook with a-priori codes. Two study team members (A.S. and N.W.) read all transcripts, conducted data familiarization, and open-coded a subset of transcripts to identify codes and preliminary themes. Research team members from Zambia and Viet Nam who participated in data collection, alongside their debrief notes, further informed additional codes and themes. The codebook was iteratively refined during the initial data familiarization and coding process, with inductive codes added along with themes. A.S. and N.W. initially double-coded a subset of six transcripts, compared and discussed coding and interpretation of codes and preliminary themes. When the codebook was finalized, all codes were systematically applied across all the transcripts.

Following this process, A.S., who has experience with qualitative research and analysis, coordinated with data collectors to discuss interpretation and conducted primary coding. N.W., an expert in qualitative methods and analysis, oversaw the analytic process. Upon completion of coding, the data were organized into a framework/chart format for key themes that included summaries of findings. Themes were iteratively compared and explored across participant types and sub-categories (participant type, age, sex, TB status) to facilitate the identification.

Analysts maintained reflexive memos and analytic memos to document their thinking and decision-making throughout the analytic process. Analysts also met with data collectors and investigators in both countries throughout the data collection and analytic process to discuss and receive feedback on findings and analytic approaches.

## 3. Results

### 3.1 Descriptive analysis of participants

The sample included 76 participants (n = 40 adults undergoing TB evaluation, n = 21 healthcare workers, and n = 15 caregivers) at six primary health centers in Viet Nam and Zambia.

A demographic summary of participants is presented in Table 1. In both Viet Nam and Zambia, most healthcare workers and all caregivers identified as female (n = 36, 94%). In Viet Nam, all adults accessing TB care (n = 19, 100%) consented to both tongue swab and sputum collection, while specimen provision varied in Zambia (n = 21; 67% [n=14] consented to both tongue swab and sputum collection, 19% [n=4] to tongue swab collection only, and 14% [n=3] to sputum collection only). Among individuals recruited for the study, two males in Viet Nam declined participation due to a lack of time. All individuals recruited in Zambia agreed to participate.

**Table 1:** Demographic profile of participants (n=76) in Viet Nam and Zambia.

| **Viet Nam (n = 31)** |  |  | **Zambia (n=45)** |  |  |
| --- | --- | --- | --- | --- | --- |
| Healthcare workers, n = 7 |  |  | Healthcare workers, n = 14 |  |  |
|  | Male | 1 (14%) |  | Male | 1 (7%) |
|  | Female | 6 (86%) |  | Female | 13 (93%) |
| Adults with presumptive TB, n = 19 |  |  | Adults with presumptive TB, n = 21 |  |  |
|  | Male | 10 (57%) |  | Male | 9 (43%) |
|  | Female | 9 (43%) |  | Female | 12 (57%) |
|  | Swab + Sputum | 19 (100%) |  | Swab + Sputum | 14 (67%) |
|  | Swab only | 0 (0%) |  | Swab only | 4 (19%) |
|  | Sputum only | 0 (0%) |  | Sputum only | 3 (14%) |
| Caregivers, n = 5 |  |  | Caregivers, n = 10 |  |  |
|  | Male | 0 (0%) |  | Male | 0 (0%) |
|  | Female | 5 (100%) |  | Female | 10 (100%) |
|  | Swab + Sputum | 2 (40%) |  | Swab + Sputum | 4 (40%) |
|  | Swab only | 1 (20%) |  | Swab only | 4 (40%) |
|  | Sputum only | 0 (0%) |  | Sputum only | 1 (10%) |
|  | Swab and Stool | 2 (40%) |  | Stool only | 1 (10%) |

**Table 2:** Themes affecting acceptability and feasibility of tongue swabs based on stakeholder groups.

|  | Stakeholders |  |  |
| --- | --- | --- | --- |
|  | People accessing TB services | Caregivers | Health workers |
| **Acceptability** |  |  |  |
| *How willing individuals or groups are to receive or deliver a healthcare service, treatment, or program based on anticipated or experienced cognitive and emotional responses to it (Sekhon et al., 2017).* | Ease of collection<br>Person-centeredness of sampling<br>Readiness to use<br>Unknown accuracy*; lower perceived accuracy compared to sputum samples<br>Familiarity with sputum sampling methods in TB; lack of familiarity with tongue swabs to diagnose TB |  |  |
|  | Increased comfort during sample collection compared to sputum methods<br>Familiarity due to COVID-19 nasal swabs |  | Potential increased risk of TB transmission to HCWs |
| **Feasibility** |  |  |  |
| *The extent to which an approach can be successfully implemented and integrated within a given context, setting, or healthcare system. It assesses whether an intervention is practical, realistic, and viable to deliver under real-world conditions (Eldridge et al., 2016).* | Speed of sample collection (e.g. few minutes vs. ~30 minutes for sputum for those who had difficulty to expectorate sputum) |  |  |
|  | Physical comfort during sampling compared to physical discomfort during sputum collection | Ease of use for children compared to sputum collection | Potential to significantly increase diagnostic yield (e.g. test children, people who cannot expectorate)<br>Increased hygiene for HCWs handling tongue swabs over sputum bottles |
| **Process of implementation** |  |  |  |
| *The steps and activities involved in putting an intervention into practice (Damschroder et al., 2009).* | Engagement and support (e.g. health education, counseling)<br>Understand test accuracy prior to testing |  |  |
|  | Potential for self-swabbing or HCW-directed swabbing |  | Develop standardized information on swab's time to result, collection procedures, and accuracy<br>Ensure adequate PPE is provided to HCWs |

\*Accuracy is a theme that significantly influenced test acceptability and preference

### 3.2 Themes

Tongue swabs were generally considered feasible and easy to collect, with many participants welcoming them as a complementary or alternative approach to sputum-based testing. Many participants across groups—people with presumptive TB, healthcare workers, and caregivers—mentioned usability, diagnostic accuracy, hygiene, reduced transmission risks, and trust as key considerations influencing tongue swab acceptability (Figure 1). Nearly every person seeking TB care, healthcare worker, and caregiver in Viet Nam, and most participants in Zambia, preferred tongue swab collection over sputum collection. Overall, all participants in Viet Nam and most in Zambia indicated that tongue swabs were acceptable.

**Figure 1.**
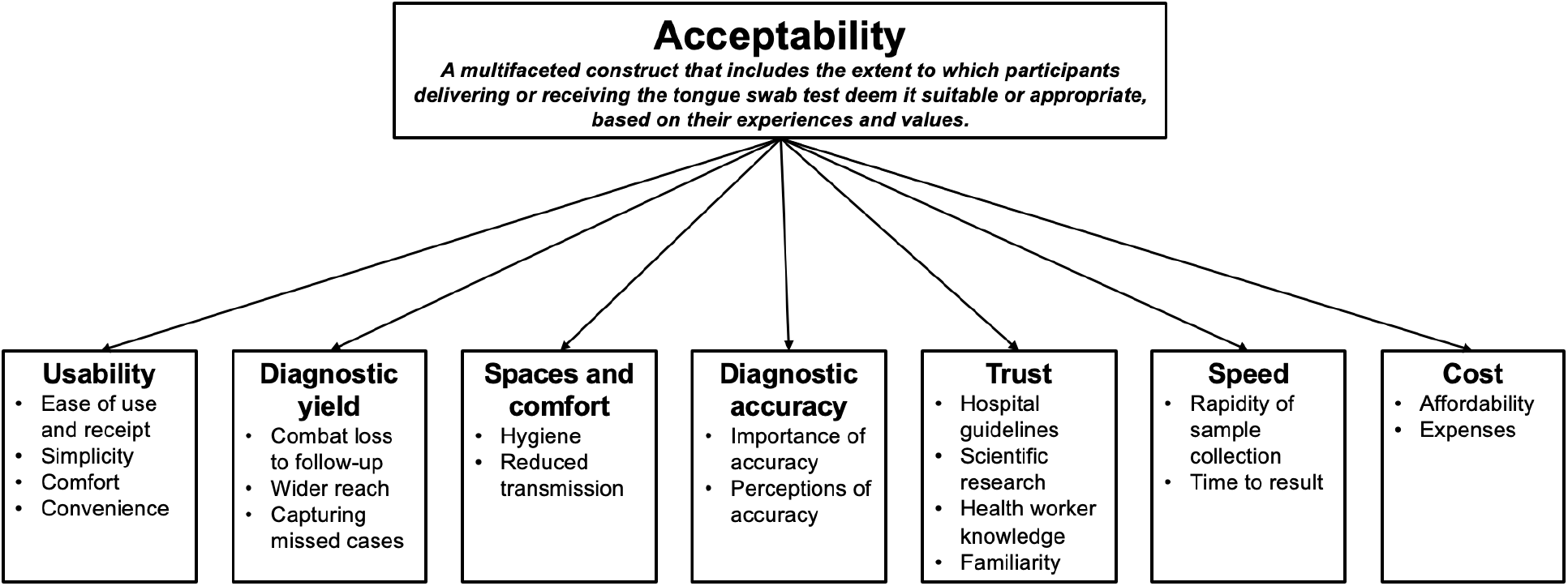
Domains of acceptability for tongue swabs adapted from Martin et al. (2022); Sekhon et al. (2017)

#### 3.2.1 Usability

Usability from the healthcare worker and caregiver perspectives drove acceptability of and preference for tongue swabs. Many individuals accessing TB care, healthcare workers, and caregivers discussed the ease of use and receipt of tongue swabs. All participants in Viet Nam and most in Zambia highlighted the advantages of a tongue swab, including convenience, simplicity, and comfort during collection. Several participants also compared the methods for collecting tongue swabs to sputum, calling the tongue swab “simpler,” “easier,” “faster,” and “gentler.” Many people accessing TB care described difficulty in expectorating sputum, even when they attempted to produce sputum on different occasions. Some participants highlighted the acceptability of tongue swabs for various target groups:

> *“It [tongue swab] would just encourage me because it is very much easier to use, it is user-friendly, such that even if a person is very ill, they could not fail to collect a sample*.*”* – Zambia, Adult undergoing TB evaluation

Many participants in Viet Nam and the majority of those in Zambia noted that producing sputum was not possible for everyone. All participant groups, especially caregivers and healthcare workers, emphasized the benefits of the tongue swab to test children, older adults, and those who struggled to expectorate.

Caregivers frequently described the ease of tongue swab sampling for their children, many of whom struggled to produce a quality sputum sample. Healthcare workers also often noted how tongue swabs made TB testing more accessible for participants with dry coughs and discussed the potential for tongue swabs to diagnose more children with TB.

> *“We are having low numbers in our children below the age of five…Because for them, producing sputum, they swallow it. When it comes to swabbing, we are able to do it…So, tongue swab is easy. We are able to collect the sample that you want, unlike them producing sputum, which when they cough out, you are able to tell that there is sputum, but they would cough it out and swallow it again…And in such cases, we are finding it difficult, hence we have low numbers of children with actually confirmed TB compared to adults*.*”* – Zambia, Healthcare worker

#### 3.2.2 Diagnostic yield

Healthcare workers in both countries noted that tongue swabs could increase diagnostic yield. For the participants who failed to expectorate during their first clinic visit, healthcare workers often provided those accessing TB diagnostic services with bottles for home collection and instructed them to return to the clinic after completing sputum collection. Healthcare workers perceived tongue swabs as a more convenient and user-friendly alternative to requiring patients to return to the clinic on a subsequent day.

> *“The [swab] will be much more convenient because we will definitely not have any cases where we have to schedule an appointment for the next day to wait from the time the person goes to sleep until they wake up to try to collect sputum samples. And for tongue swab samples, we will also collect them right at the examination session*.*”* – Viet Nam, Healthcare worker

Healthcare workers explained the effectiveness of tongue swabs in reaching communities that might otherwise be missed, including individuals who are reluctant to associate themselves with TB and may not produce sputum. Often, healthcare workers in both Viet Nam and Zambia described how the quality of a sample can influence its accuracy. A healthcare worker in Viet Nam described how tongue swabs meet quality requirements compared to sputum:

> *“Between the sputum sample and the tongue swab*…*if we do not talk about the accuracy, I find the tongue swab more effective than sputum. The percentage of quality tongue swabs is higher than that of sputum…The patient may have difficulty to produce sputum or the patients do not obtain it correctly…As for the tongue swab, I think that all the tongue swabs I have collected meet the quality requirements*.*”* – Viet Nam, Healthcare worker

Delayed treatment, often from inconvenient sampling methods like sputum-based testing, was felt to lead to more TB deaths:

> *“Maybe I will say if someone coughs out saliva only, because I know if you cough out saliva only, we might miss the patients with TB…Meanwhile, that patient has TB but is unable to cough out the sputum, which is meant for it. So, we are missing a lot of cases, actually…It usually leads to a lot of TB deaths. We have the X-ray machine, but there is no one working there at the moment, we don’t have any personnel, so some even fail, some don’t even have money to go for X-ray, as a result, we will be having a lot of deaths due to delaying of starting treatment*.*”* – Zambia, Healthcare worker

#### 3.2.3 Spaces and comfort

Comfort during specimen collection, including hygiene and transmission risk considerations, informed participants’ attitudes toward tongue swab-based TB testing. Many participants in both countries expressed that the tongue swab was comfortable, although more individuals in Zambia described mild side effects (e.g., nausea, itchiness). Around half of the adults undergoing TB evaluation in Zambia mentioned pain during the sputum expectoration process, compared to the few individuals in Viet Nam who reported discomfort.

Hygiene and lower risk of transmission, described by all participant groups, drove the acceptability of tongue swabs, as explained by a participant:

> *“Although there wasn’t a separate area for expectoration here, I had to go to the middle of the park and spit sputum out, which was a bit inconvenient and uncivilized [laughing]. It was uncivilized…Suddenly, I went to the middle of the park and spitted sputum out while there were a lot of people there*.*”* – Viet Nam, Adult undergoing TB evaluation

People accessing TB care generally felt that collecting tongue swabs posed a lower risk of TB transmission. One individual from Viet Nam said he prefers the tongue swab because “*That’s the best way to do it [test] without infecting everyone around me*.*”* – Viet Nam, Adult undergoing TB evaluation

However, healthcare workers expressed mixed feelings towards the risk of TB transmission in collecting tongue swabs, with some healthcare workers expressing fear that adults and children may cough during tongue swab collection, posing a greater danger to the safety of healthcare workers:

> *“Fears when conducting a tongue swab are there because if they open their mouth and you are swabbing their tongue, they might start coughing right there. Since you are near them when swabbing their tongue, they might cough on your hands and on you…there is need for protective measures”* – Zambia, Healthcare worker

Many healthcare workers believed that the tongue swab was safer for communities themselves, especially if they wore protective equipment, such as masks:

> *“If I take a sample from the throat, the risk for healthcare workers may be higher. That’s because I directly take it*..*As for the sputum sample, patients go to a corner and expectorate…but it can also lead to community transmission because although it may not infect healthcare workers, others passing by, for example, have a higher chance of getting infected*…*Taking a tongue swab sample may also cause a higher risk for healthcare workers. However, if we protect ourselves well*,…*I don’t think the risk is very high*.*”* – Viet Nam, Healthcare worker

#### 3.2.4 Diagnostic accuracy

##### 3.2.4.1 Importance of accuracy

Most participants accessing TB care, healthcare workers, and caregivers agreed that an accurate diagnosis was vital to their trust in tongue swabs. In both Viet Nam and Zambia, many adults with presumptive TB discussed their preference for undergoing a more accurate but more difficult diagnostic test, compared to an easier but less accurate one. The desire to detect and cure one’s disease largely shaped participants’ perceptions of the tongue swab.

> *“It’s true that the tongue swab test is hygienic and fast, but it’s not reassuring. I’m the one who wish to detect the disease, I’m not scared of being painful. I just want to do everything to detect my disease accurately*.*”* – Viet Nam, Adult undergoing TB evaluation

Most adult participants seeking care in both countries prioritized diagnostic accuracy over usability.

> *“I would pick the one [test] which is able to detect the disease*…*Because of the need to get better…So if it happens that I pick something easy but not able to detect TB, then there is absolutely nothing I am doing*.*” –* Zambia, Adult undergoing TB evaluation

Every healthcare worker in Viet Nam and Zambia discussed the importance of choosing a TB test with high diagnostic accuracy. Although many healthcare workers noted that the tongue swab was easier to collect, a lack of knowledge about its accuracy posed barriers to the acceptability of tongue swab-based tests. It sparked healthcare workers’ “concern about the results.” For instance, when asked whether healthcare workers trusted the tongue swab, one noted:

> *“This [test] does not have any results yet. I don’t know. I cannot evaluate it yet because, for example, with that [sputum] sample, I already know what the patient’s results are and whether they are accurate or not. I do not know if the [swab] is as accurate as a sputum sample*.*”* – Viet Nam, Healthcare worker

##### 3.2.4.2 Perceptions of Accuracy

When discussing the perceived accuracy of tongue swabs, participants expressed mixed opinions on the fluid locations within the body that indicated the accuracy of test samples. In Viet Nam, some believed in the accuracy of the tongue swab because “sputum passes through the throat,” while a number of participants felt that sputum itself would be more accurate. In both countries, several individuals accessing TB care, healthcare workers, and caregivers discussed the lungs, rather than the tongue, as the source of TB and a more accurate indicator of TB diagnosis:

> *“[Compared to the tongue swab], I like the sputum sample because it comes from the chest, and from my chest is where everything I take, even if someone is smoking, is drinking beers, is doing whatever, something which can give someone else TB. They are coming from the chest, and that is where the sputum comes from*…*I mean to say because even in the chest itself you can find the signs of TB. And the sputum comes from the chest*.*”* – Zambia, Adult undergoing TB evaluation

#### 3.2.5 Trust

##### 3.2.5.1 The health system

Hospital guidelines, scientific research, and healthcare worker knowledge all influenced trust in the tongue swab, particularly among people accessing TB services and caregivers. One adult accessing care in Zambia discussed how her trust in research informs her trust in the tongue swab:

> *“I am sure the researchers have researched and they have seen that that it can be trusted because they, I’m sure they have already run this, they can’t bring it here when they have not tested on it anywhere else, so I’m sure for them to bring it here I’m confident that it is okay*.*”* – Zambia, Adult undergoing TB evaluation

Another adult accessing TB care in Zambia explained why she trusts the tongue swab and her reasons for following doctors’ recommendations:

> *“What has made me trust it [tongue swab] that it’s effective, is simply because the doctors don’t lie, so I believe that it is also a method of testing for TB, it’s not fake because we survive through the hospitals. [Doctors don’t lie] because they are educated, they learn about us, the same human beings, and know how to diagnose TB*…*I wouldn’t refuse tongue swabs because I have to follow the doctor’s instructions…I want to get healed, I need help, so I should follow what the health facility says*.*”* – Zambia, Adult undergoing TB evaluation

Compared to those in Zambia, more people accessing TB care in Viet Nam emphasized their complete trust in doctors and healthcare workers to guide their health decision-making. In both countries, individuals seeking care generally described healthcare workers as knowledgeable and thorough in their recommendations and described hospitals as places of help and healing.

> *“As for the perspective of ensuring the safety and everything, the medical field will have guidance when introducing the sample. They will have considered every aspect. To make it work. That sample. Whether it’s popular or easy to use*.*”* – Viet Nam, Adult undergoing TB evaluation

##### 3.2.5.2 Familiarity

Familiarity with sputum-based testing and COVID-19 nasal swabs also influenced trust in TB diagnostic tests. For a person seeking TB care in Zambia, a lack of knowledge and familiarity with tongue swabs drove his preference for sputum tests:

> *“We are used to the bottles*.. *Yes, so we have never heard of these new test kits [for the tongue]*… *We have never tried them out…So that is why I preferred the bottle*.*”* – Zambia, Adult undergoing TB evaluation

People accessing TB care, caregivers, and healthcare workers in both Viet Nam and Zambia described positive and negative associations with COVID-19 swabs that affected their perceptions of tongue swabs. While overall preferred as compared to sputum, some participants, particularly caregivers of children, had hesitancy using a swab based on experience with COVID-19 swab testing. A healthcare worker in Viet Nam discussed misperceptions of the tongue swab procedure and the evolving attitudes of those accessing TB services:

> *“Initially, when comparing it with COVID sample collection, everyone thought it would involve deep sampling. But when we mentioned that it was just a small swab on the tongue, and after experiencing it, everyone preferred it over sputum collection. However, there were a few cases where patients could expectorate on their own and didn’t want to put anything in their mouth. I think the majority of people would prefer the tongue swab test because of its simple procedure. Anyone can provide the sample*.*”* – Viet Nam, Healthcare worker

#### 3.2.6 Speed

Nearly all participants in Viet Nam and several in Zambia mentioned the quick speed of collecting tongue swabs. Several participants also mentioned speed as a factor that drove their acceptability of and preference for tongue swab-compared to sputum-based testing.

In both countries, several participants cited the importance of a rapid turnaround time for receiving results because of the belief that an earlier diagnosis would lead to earlier medication and treatment. Our interviews suggest that informing patients of the rapidity of testing results can help them determine their preferred testing method. One participant explained, *“I do not know whether the tongue swab or sputum test is better. I can just choose what is fast*.*”* – Zambia, Adult undergoing TB evaluation

#### 3.2.7 Cost

A few individuals in Viet Nam noted how costs factored into their perceptions of TB diagnostic tests. One caregiver in Viet Nam said, *“If we are infected, it’ll cost a fortune to treat the disease*.*”* When comparing the tongue swab and sputum-based tests, an individual noted, *“I want to know about and compare the expenses for these tests in the future. If the difference between the expenses of both methods is suitable to me, I would definitely choose the new and faster method*.*”* – Viet Nam, Adult undergoing TB evaluation

Expenses were not only a concern for those seeking TB care, but also for individuals working within the health system. Due to the individuals who are unable to expectorate or produce a quality sample, a healthcare worker described the potential costs associated with sputum-based tests; *“many people could not produce quality sputum, so I had to guide them again, which would lead to an issue with the consumable materials that came with repeating sputum collection, definitely costing more money*.*”* – Viet Nam, Healthcare worker

#### 3.2.8 Acceptability and Feasibility

To better understand tongue swab uptake during our study and its potential for widespread adoption, findings were categorized under the domains of acceptability and feasibility.

## 4. Discussion

Our study is among the first to evaluate tongue swab usability, acceptability, and preferences for people undergoing TB evaluation, caregivers, and healthcare workers. Most participants generally found tongue swabs acceptable, noting the ease of sample collection. High ease of tongue swab collection allowed participants who struggled to produce sputum, such as children and individuals unable to expectorate, to provide a sample for testing. Among the participant groups, healthcare workers most frequently described the ability of tongue swabs to increase diagnostic yield. Other drivers of tongue swab acceptability included hygiene and transmission risk considerations, trust in hospital guidelines, the research required for test rollout, and healthcare worker knowledge. Nearly all participants considered test accuracy in their perceptions of tongue swabs. Our findings suggest that broader implementation of tongue swabs can facilitate more convenient and comfortable testing experiences and engage more individuals in the TB diagnostic and evaluation cascade.

The limitations of sputum-based tests are well-documented. Many individuals, including children and people living with HIV, have difficulty producing or are unable to produce sputum altogether (Andama et al., 2022; Broger et al., 2023; Church et al., 2024; Codsi et al., 2023). Nearly all participants in our study preferred the tongue swab to the sputum test, supporting findings from prior studies utilizing quantitative preference elicitation methods (Castro et al., 2024; Shah et al., 2025). Qualitative data from healthcare workers also demonstrated a preference for tongue swabbing over sputum collection (Codsi et al., 2023).

Several healthcare workers also described their aversion to handling sputum samples compared to the more hygienic tongue swab. Many healthcare workers also perceived that the general risk of TB transmission was lower when using tongue swabs compared to sputum tests and that their own risk of transmission was low when using protective equipment, primarily high-quality N95 masks. Our findings align with a study that explored the preferences of South African healthcare workers in using tongue swabs to diagnose TB (Codsi et al., 2023). These healthcare workers generally preferred tongue swabs over sputum-based tests, described an aversion to sputum, and perceived a low risk of TB exposure due to mask protection. In our study, however, several healthcare workers and people accessing TB care raised concerns about TB transmission when using tongue swabs; they described how sample collection using swabs requires proximity between healthcare workers and people with symptoms of TB, whereas sputum expectoration typically occurs in a more isolated area away from healthcare workers. The healthcare workers who spoke of this difference also noted their confidence in masks to limit their TB exposure, similar to the statements by healthcare workers in Codsi et al.’s (2023) study. Given the lack of qualitative data in TB diagnostics, we draw upon HIV self-testing (HIVST) literature. Similar to TB tests, HIV self-tests offer a rapid, oral test as an alternative to blood-based tests, the primary method of testing. Overall, the domains of tongue swab acceptability expressed by our study participants align with an HIVST study that evaluated participants’ attitudes toward oral and blood-based tests (Mantell et al., 2022). Themes that emerged in both studies include perceived rapidity of test results, lack of pain or discomfort, ease of use, and trust in results and healthcare workers.

Many participants noted the importance of diagnostic accuracy, rapid result availability, and diagnostic yield when considering tongue swab acceptability. Several of our participants described a belief that sputum-based tests provided more accurate results compared to tongue swab-based tests because TB is found in the lungs. Similarly, several participants in an HIVST study believed that blood-based, rather than saliva-based, tests could provide more accurate results because HIV is present in the blood (Bwalya et al., 2020). In a multicountry discrete choice experiment, the TB diagnostic tests’ most strongly valued features were high diagnostic accuracy, fast time to result, and free cost; sample type was the least important feature (Shah et al., 2025).

Our study suggests that the current WHO guidelines on TB diagnostics could expand to encourage tongue swab tests. Many participants in our study discussed accuracy as a notable consideration for accepting tongue swab-based TB testing. Tongue swabs should be prioritized in settings without TB testing or where only sputum smear microscopy is offered. Compared to sputum smear microscopy, tongue swab-based molecular testing provides greater sensitivity (Steadman et al., 2025) and is likely to meet the minimum accuracy thresholds for a non-sputum rapid diagnostic test (Steadman et al., 2025). In settings where sputum-based molecular testing is provided, tongue swabs should be offered as an additional service, particularly for people unable to expectorate sputum.

Diagnostic accuracy alone does not determine test preference. Diagnostic yield has been suggested as an important metric for assessing TB diagnostic tests (Broger et al., 2023). Within COVID-19 testing, nasopharyngeal swabs also exhibited lower average viral loads than sputum, yet these swabs were preferred for detecting SARS-CoV-2 and improved diagnostic yield (Yerlikaya et al., 2024). Most of our study participants also preferred tongue swabs over sputum tests, and many healthcare workers preferred tongue swabs because of the potential to increase diagnostic yield and capture more TB cases. Implementing accurate TB tests that emphasize diagnostic yield can target disadvantaged and vulnerable populations. Our findings show that the feasibility and acceptability of tongue swabs can allow further diagnostic yield compared to sputum-based molecular testing alone. Many participants in our study who consented to sputum-based tests had difficulty with or could not produce sputum, but could use the tongue swab. In many high TB burden contexts, those who are not able to produce sputum are asked to return to the clinic at a later time, and/or take a sputum collection tube home to return with a sample (Datta et al., 2017). This approach risks more costs to people with symptoms of TB, loss to follow-up, delays in diagnosis and treatment, and disease transmission and progression. In addition to adults, adolescents and children are at an increased risk of not returning for diagnosis (Pala et al., 2018). Tongue swabs available for those who cannot produce sputum offer an alternative to mitigate this pressing challenge and avert missed diagnoses (Bailey et al., 2011).

Several aspects must be considered to facilitate effective implementation. First, the risk of TB transmission to healthcare workers must be mitigated. Adequate protective equipment, such as N95 masks, must be provided to all healthcare workers collecting tongue swabs. Providing isolated, open areas for people to swab can also limit potential exposure to TB bacteria. Educational tools that teach people about TB transmission risks, discourage coughing during sample collection, or demonstrate how to self-swab can further enhance the safety of healthcare workers. The high usability of tongue swabs offers the potential for self-testing as a possible diagnostic method (Codsi et al., 2023), as observed through COVID-19 self-collected nasal swabs (Yerlikaya et al., 2024). Second, measures to build trust are critical to familiarize people with the novel TB tongue swabs. Several of our participants distrusted the tongue swab because they lacked an understanding of its swabbing method and purpose. Drawing upon HIVST, educating participants before testing is a critical strategy that can promote acceptance and dispel fear (Mukora-Mutseyekwa et al., 2022). Showing educational videos or distributing pamphlets that provide more information on tongue swabs can increase comfort and acceptability. Within COVID-19 testing, leveraging community healthcare workers as trusted messengers was used to expand diagnostic testing into communities (Stadnick et al., 2023; Ziolkowski et al., 2024), and can be employed for tongue swabs. Other trusted messengers facilitating HIVST include peers and known contacts (Lewis-Kulzer et al., 2024), community-based distribution agents (Harrison et al., 2022), and village healthcare workers (Amstutz et al., 2020). Leveraging similar stakeholders can facilitate tongue swab implementation. Third, informing participants of test accuracy and result turnaround time can encourage consent to swabbing.

Our study had several limitations. First, the study was conducted in only urban areas in Viet Nam; the data was collected in three health centers in the same city, which may limit the generalizability of our findings. However, since the prevalence of TB is significantly higher in urban areas in Viet Nam (Nguyen et al., 2020) and other high burden settings (Lei et al., 2023), our study focused on highly impacted populations. Further, the health centers in Zambia (urban, peri-urban, and rural health centers) represented a diverse range of settings. Second, we used interviews with caregivers as proxies to understand children’s experiences with TB diagnostic tests. This limited learning directly from children; however, it may have allowed for more precise and thorough discussions based on caregivers’ experiences from direct participation in the TB evaluation consultation and observation of the sample collection process. Third, at the time of the interviews, participants had yet to receive their test results and were unsure of the accuracy of tongue swabs compared to sputum-based tests. This limited some participants’ ability to establish their preference for and acceptance of tongue swabs. However, conducting interviews right after the sample collection helped combat recall bias, and reflects preferences independent of accuracy data.

## 5. Conclusion

Our findings suggest that tongue swabs are generally an acceptable and preferred method for TB testing. Tongue swabs, considered highly feasible and easy to use, demonstrate the potential to improve TB diagnostic evaluation in high-burden settings. Favorable aspects of the tongue swab included high usability, increased diagnostic yield, improved hygiene, and faster speed to test. Integrating tongue swabs into TB diagnostics is critical to reach a broader population. Tongue swabs offer a more convenient and user-friendly method to evaluate groups that may have difficulty producing sputum and often are clinically diagnosed with TB, such as children. Future facility- and community-based promotion of tongue swab collection should target drivers of acceptability by developing and including standardized information on the swab’s time to result, collection procedures, and accuracy compared to sputum-based tests. Context-specific considerations for personal protective wear for swab collectors and the physical location of tongue swab collection may minimize discomfort and transmission concerns. Given that the acceptability of tongue swabs and potential barriers to adoption and use varied between countries and participants, more research is needed to understand country- and healthcare system-specific implementation strategies. Finally, further studies on community-based and self-collection of tongue swabs are required to fully understand this alternative diagnostic method’s potential.

## Data Availability

Qualitative data from this study are not publicly available to protect potential identification of participants based on their detailed responses.

## References

Amstutz, A., Kopo, M., Lejone, T. I., Khesa, L., Kao, M., Muhairwe, J., Glass, T. R., & Labhardt, N. D. (2020). “If it is left, it becomes easy for me to get tested”: Use of oral self-tests and community health workers to maximize the potential of home-based HIV testing among adolescents in Lesotho. Journal of the International AIDS Society, 23 Suppl 5(Suppl 5), e25563. 10.1002/jia2.25563

Andama, A., Whitman, G. R., Crowder, R., Reza, T. F., Jaganath, D., Mulondo, J., Nalugwa, T. K., Semitala, F. C., Worodria, W., Cook, C., Wood, R. C., Weigel, K. M., Olson, A. M., Lohmiller Shaw, J., Kato-Maeda, M., Denkinger, C. M., Nahid, P., Cangelosi, G. A., & Cattamanchi, A. (2022). Accuracy of Tongue Swab Testing Using Xpert MTB-RIF Ultra for Tuberculosis Diagnosis. Journal of Clinical Microbiology, 60(7), e00421–22. 10.1128/jcm.00421-22

Bailey, S. L., Roper, M. H., Huayta, M., Trejos, N., López Alarcón, V., & Moore, D. a. J. (2011). Missed opportunities for tuberculosis diagnosis. The International Journal of Tuberculosis and Lung Disease: The Official Journal of the International Union Against Tuberculosis and Lung Disease, 15(2), 205–210, i.

Broger, T., Koeppel, L., Huerga, H., Miller, P., Gupta-Wright, A., Blanc, F.-X., Esmail, A., Reeve, B. W. P., Floridia, M., Kerkhoff, A. D., Ciccacci, F., Kasaro, M. P., Thit, S. S., Bastard, M., Ferlazzo, G., Yoon, C., Van Hoving, D. J., Sossen, B., García, J. I., … Fielding, K. (2023). Diagnostic yield of urine lipoarabinomannan and sputum tuberculosis tests in people living with HIV: A systematic review and meta-analysis of individual participant data. The Lancet Global Health, 11(6), e903–e916. 10.1016/S2214-109X(23)00135-3

Bwalya, C., Simwinga, M., Hensen, B., Gwanu, L., Hang’andu, A., Mulubwa, C., Phiri, M., Hayes, R., Fidler, S., Mwinga, A., Ayles, H., Bond, V., & the HPTN 071 (PopART) study team. (2020). Social response to the delivery of HIV self-testing in households: Experiences from four Zambian HPTN 071 (PopART) urban communities. AIDS Research and Therapy, 17(1), 32. 10.1186/s12981-020-00287-y

Castro, M. del M., Le, H., Muzazu, S., Pham, N., Trinh, T., Nyirenda, H., Shabalu, P., West, N., Phan, H., Cattamanchi, A., Denkinger, C. M., Muyoyeta, M., & Kerkhoff, A. D. (2024). Preferences for tongue swab-versus sputum-based testing in the context of TB care: A Best-Worst Scaling exercise in Vietnam and Zambia. Infectious Diseases (except HIV/AIDS). 10.1101/2024.12.22.24319450

Church, E. C., Steingart, K. R., Cangelosi, G. A., Ruhwald, M., Kohli, M., & Shapiro, A. E. (2024). Oral swabs with a rapid molecular diagnostic test for pulmonary tuberculosis in adults and children: A systematic review. The Lancet Global Health, 12(1), e45–e54. 10.1016/S2214-109X(23)00469-2

Codsi, R., Errett, N. A., Luabeya, A. K., Van As, D., Hatherill, M., Shapiro, A. E., Lochner, K. A., Vingino, A. R., Kohn, M. J., & Cangelosi, G. A. (2023). Preferences of healthcare workers using tongue swabs for tuberculosis diagnosis during COVID-19. PLOS Global Public Health, 3(9), e0001430. 10.1371/journal.pgph.0001430

Damschroder, L. J., Aron, D. C., Keith, R. E., Kirsh, S. R., Alexander, J. A., & Lowery, J. C. (2009). Fostering implementation of health services research findings into practice: A consolidated framework for advancing implementation science. Implementation Science, 4(1), 50. 10.1186/1748-5908-4-50

Datta, S., Shah, L., Gilman, R. H., & Evans, C. A. (2017). Comparison of sputum collection methods for tuberculosis diagnosis: A systematic review and pairwise and network meta-analysis. The Lancet Global Health, 5(8), e760–e771. 10.1016/S2214-109X(17)30201-2

Eldridge, S. M., Lancaster, G. A., Campbell, M. J., Thabane, L., Hopewell, S., Coleman, C. L., & Bond, C. M. (2016). Defining Feasibility and Pilot Studies in Preparation for Randomised Controlled Trials: Development of a Conceptual Framework. PLOS ONE, 11(3), e0150205. 10.1371/journal.pone.0150205

Gale, N. K., Heath, G., Cameron, E., Rashid, S., & Redwood, S. (2013). Using the framework method for the analysis of qualitative data in multi-disciplinary health research. BMC Medical Research Methodology, 13(1), 117. 10.1186/1471-2288-13-117

Global Tuberculosis Report 2024. (2024). World Health Organization.

Harrison, L., Kumwenda, M., Nyirenda, L., Chilongosi, R., Corbett, E., Hatzold, K., Johnson, C., Simwinga, M., Desmond, N., & Taegtmeyer, M. (2022). “You have a self-testing method that preserves privacy so how come you cannot give us treatment that does too?” Exploring the reasoning among young people about linkage to prevention, care and treatment after HIV self-testing in Southern Malawi. BMC Infectious Diseases, 22(S1), 395. 10.1186/s12879-022-07231-7

High priority target product profiles for new tuberculosis diagnostics: Report of a consensus meeting (p. 98). (2014). World Health Organization. https://www.who.int/publications/i/item/WHO-HTM-TB-2014.18

Lei, Y., Wang, J., Wang, Y., & Xu, C. (2023). Geographical evolutionary pathway of global tuberculosis incidence trends. BMC Public Health, 23(1), 755. 10.1186/s12889-023-15553-7

Lewis-Kulzer, J., Olugo, P., Gutin, S. A., Kwena, Z. A., Nishimura, H., Thorp, M., Agot, K., Ayieko, B., Bukusi, E. A., Oluoch, L., Angawa, D., Thirumurthy, H., & Camlin, C. S. (2024). “There is no need to leave the beach to test”: A qualitative study of HIV self-testing knowledge and acceptability of HIV self-test kit distribution among social networks of fishermen in western Kenya. In Review. 10.21203/rs.3.rs-5090648/v1

Mantell, J. E., Khalifa, A., Christian, S. N., Romo, M. L., Mwai, E., George, G., Strauss, M., Govender, K., & Kelvin, E. A. (2022). Preferences, beliefs, and attitudes about oral fluid and blood-based HIV self-testing among truck drivers in Kenya choosing not to test for HIV. Frontiers in Public Health, 10, 911932. 10.3389/fpubh.2022.911932

Martin, S., Parlier-Ahmad, A. B., Eglovitch, M., Ondersma, S. J., Svikis, D. S., & Martin, C. E. (2022). Project BETTER: Preliminary Feasibility and Acceptability of a Technology-Delivered Educational Program for Pregnant and Postpartum People with Opioid Use Disorder. Women’s Health Reports, 3(1), 834–843. 10.1089/whr.2022.0046

Mukora-Mutseyekwa, F., Mundagowa, P. T., Kangwende, R. A., Murapa, T., Tirivavi, M., Mukuwapasi, W., Tozivepi, S. N., Uzande, C., Mutibura, Q., Chadambuka, E. M., & Machinga, M. (2022). Implementation of a campus-based and peer-delivered HIV self-testing intervention to improve the uptake of HIV testing services among university students in Zimbabwe: The SAYS initiative. BMC Health Services Research, 22(1), 222. 10.1186/s12913-022-07622-1

Nguyen, H. V., Tiemersma, E. W., Nguyen, H. B., Cobelens, F. G. J., Finlay, A., Glaziou, P., Dao, C. H., Mirtskhulava, V., Nguyen, H. V., Pham, H. T. T., Khieu, N. T. T., De Haas, P., Do, N. H., Nguyen, P. D., Cung, C. V., & Nguyen, N. V. (2020). The second national tuberculosis prevalence survey in Vietnam. PLOS ONE, 15(4), e0232142. 10.1371/journal.pone.0232142

Pala, S., Bhattacharya, H., Lynrah, K., Sarkar, A., Boro, P., & Medhi, G. (2018). Loss to follow up during diagnosis of presumptive pulmonary tuberculosis at a tertiary care hospital. Journal of Family Medicine and Primary Care, 7(5), 942. 10.4103/jfmpc.jfmpc_161_18

Regati, M., Srikanth, E., & Gowrinath, K. (2021). Study of quality of sputum being submitted for smear examination. Journal of Clinical and Scientific Research, 10(3), 145–150. 10.4103/jcsr.jcsr_25_21

Sakundarno, M., Nurjazuli, N., Jati, S. P., Sariningdyah, R., Purwadi, S., Alisjahbana, B., & van der Werf, M. J. (2009). Insufficient quality of sputum submitted for tuberculosis diagnosis and associated factors, in Klaten district, Indonesia. BMC Pulmonary Medicine, 9, 16. 10.1186/1471-2466-9-16

Sandelowski, M. (1995). Sample size in qualitative research. Research in Nursing & Health, 18(2), 179–183. 10.1002/nur.4770180211

Sekhon, M., Cartwright, M., & Francis, J. J. (2017). Acceptability of healthcare interventions: An overview of reviews and development of a theoretical framework. BMC Health Services Research, 17(1), 88. 10.1186/s12913-017-2031-8

Shah, K., Nalugwa, T., Marcelo, D., Nakawunde, R., Trinh, T., Shankar, D., Nakaweesa, A., Schraufnagel, A., Andama, A., Christopher, D., Van Luong, D., Theron, G., Worodria, W., Del Mar Castro, M., Nahid, P., Denkinger, C. M., Cattamanchi, A., Yu, C., & Kerkhoff, A. D. (2025). A Multi-Country Evaluation of Patient Preferences for Tuberculosis Diagnostics: A Discrete Choice Experiment to Inform WHO Target Product Profiles. Lancet Primary Care. In press. 10.1101/2024.06.19.24309124

Stadnick, N. A., Laurent, L. C., Cain, K. L., Seifert, M., Burola, M. L., Salgin, L., Watson, P., Oswald, W., Munoz, F. A., Velasquez, S. F., Smith, J. D., Zou, J., & Rabin, B. A. (2023). Community-engaged optimization of COVID-19 rapid evaluation and testing experiences: Roll-out implementation optimization trial. Implementation Science, 18(1), 46. 10.1186/s13012-023-01306-y

Steadman, A., Kumar, K. M., Asege, L., Kato-Maeda, M., Mukwatamundu, J., Shah, K., Trang, T., Ball, A., Khimani, K., Kim Dung, D. T., Michael, J. S., Christopher, D. J., Phan, H., Yerlikaya, S., Nahid, P., Denkinger, C. M., Cattamanchi, A., & Andama, A. (2025). Diagnostic accuracy of swab-based molecular tests for tuberculosis using novel near point-of-care platforms: A multi-country evaluation. Infectious Diseases (except HIV/AIDS). 10.1101/2025.04.12.25325603

Target product profile for tuberculosis diagnosis and detection of drug resistance (p. 51). (2024). World Health Organization. https://www.who.int/publications/i/item/9789240097698

Tuberculosis. (2024). World Health Organization. https://www.who.int/news-room/fact-sheets/detail/tuberculosis

Wood, R. C., Luabeya, A. K., Dragovich, R. B., Olson, A. M., Lochner, K. A., Weigel, K. M., Codsi, R., Mulenga, H., de Vos, M., Kohli, M., Penn-Nicholson, A., Hatherill, M., & Cangelosi, G. A. (2024). Diagnostic accuracy of tongue swab testing on two automated tuberculosis diagnostic platforms, Cepheid Xpert MTB/RIF Ultra and Molbio Truenat MTB Ultima. Journal of Clinical Microbiology, 62(4), e0001924. 10.1128/jcm.00019-24

Yerlikaya, S., Broger, T., Isaacs, C., Bell, D., Holtgrewe, L., Gupta-Wright, A., Nahid, P., Cattamanchi, A., & Denkinger, C. M. (2024). Blazing the trail for innovative tuberculosis diagnostics. Infection, 52(1), 29–42. 10.1007/s15010-023-02135-3

Ziolkowski, R. A., Balian, L., Sridhar, S., & Rodriguez, N. M. (2024). Improving uptake of COVID-19 testing and vaccination in a homeless population: Mixed-methods evaluation of community health worker-led education in a shelter. BMJ Open, 14(12), e087134. 10.1136/bmjopen-2024-087134

