## Supplementary Material for "“That is why I trust”: A qualitative study on acceptability and feasibility of novel tongue swab diagnostics to assess people presenting with tuberculosis symptoms in Viet Nam and Zambia"

**Appendix A. Site Locations**

| **Site locations in Viet Nam** | **Site locations in Zambia** |
| --- | --- |
| Hoang Mai | George |
| Long Bien | Ngwerere |
| Nam Tu Liem | Nakachenje |

**Appendix B. In-depth Interview Guides**

**Evaluating Acceptability and Usability for Tongue Swabs**

**Topic guide for semi-structured interviews with people testing for TB**

**Version 1.0 07 August 2023**

Background information (interviewer record information in fields below):

1. Participant ID:
2. Participant type:
3. Participant age (in years):
4. Participant sex:
5. Facility recruited from:
6. Interview location:
7. Interview date (day/month/year):
8. Name of person conducting interview:
9. Consent obtained?	Yes	No (do not proceed with interview until informed consent is obtained)

Introduction

Welcome to our interview today, we are grateful that you have volunteered to participate. We are conducting interviews with people like yourself who came to the clinic and were offered testing for TB to learn about their experiences with or thoughts about two different types of samples that are collected and used to test people for TB. Overall, the goal is to understand the acceptability, usability, and preferences for a sample for TB testing using a tongue swab or sputum which is coughing a substance from the lungs into a cup. There is no right or wrong answer. Please feel free to express your views during this interview. Your responses will be kept private and not shared with any of the staff at your local clinic – only myself and the research team will read them. Your answers will not be linked to your name and will not affect the type of treatment or care you receive. If there are any questions that you are uncomfortable answering, we can skip to the next question or stop the interview. Before we start, do you have any questions?

As a reminder, this interview will be audio recorded. I am going to start the recording now.

Warm-up Question

1. To start, please tell me about the reasons you decided to come to the clinic today.

Experiences, Perceptions, and Acceptability: Tongue Swabs

2. Today you were offered two types of testing for TB – sputum and tongue swabs. Please tell me what you think about these two types of tests.
3. Please tell me your thoughts about being tested for TB with a tongue swab. When I say tongue swab samples, I mean the swabs like this (show image – Appendix 1).

*Probe: Tell me about anything you like or dislike about tongue swabs.*

**[For participants who provided tongue swab samples]**

4. Please tell me about your experience with the tongue swab samples you provided when you were tested for TB?

*Probe: Was it easy or hard to give the tongue swab sample? Why?*

*Probe: Tell me about what you liked or disliked about the tongue swab?*

5. Please describe how you felt physically when the health worker used the tongue swab?

*Probe: Did you experience any pain or discomfort?*

6. When the health worker swabbed your tongue, please tell me what you thought about the instructions or information the health worker gave you before, during, or after collecting the sample?

*Probe: How did the health worker explain the tongue swab to you?*

*Probe: What concerns or questions did you have before, during, or after the procedure?*

*Probe: What information would you have liked to have about the tongue swab before, during, or after the procedure?*

**[For participants who provided sputum samples]**

7. Please tell me about your experience with the sputum samples?

*Probe: Was it easy or hard to give the sputum sample? Why?*

*Probe: Tell me about what you or they liked or disliked about the sputum?*

8. Please describe how you felt physically when providing the sputum sample.

*Probe: Did you experience any pain or discomfort?*

9. When the health worker collected or explained the collection of the sputum sample, please tell me what you thought about the instructions or information the health worker gave them before, during, or after collecting the sample?

*Probe: How did the health worker explain the sputum sample to you?*

*Probe: What concerns or questions did you have before, during, or after the procedure?*

*Probe: What information would you have liked to have about the sputum sample before, during, or after the procedure?*

**[For participants who did not provide tongue swab samples]**

10. Please tell me why you decided not to give a tongue swab sample today?

*Probe: What did you think when the health worker asked to give you a tongue swab?*

*Probe: Please tell me about any worries or concerns you had about having your tongue swabbed today.*

**[For all participants]**

11. Please tell me about how you would feel if you came to the clinic for TB testing and you were told you would have only a tongue swab to test for TB because it would be too hard to collect a sputum sample?

*Probe: Would having a tongue swab as your test make you feel uncomfortable or comfortable. Please tell me why.*

*Probe: Would you rather have a different test? Please tell me why.*

12. Please tell me about anything that would influence your choice to accept or decline/not accept tongue swabs for TB testing in the future?
13. Please tell me about any problems you experienced during or after the tests for TB were done that came from taking the test.

*Probe: Please describe any discomfort you experienced.*

*Probe: Please tell me about anything that was difficult or inconvenient about the test you had.*

14. Please tell me about any concerns, if any, that you have with providing a sputum sample for TB testing?
15. Please tell me about any concerns, if any, that you have with providing a tongue swab sample for TB testing?
16. If you had a choice between providing a sputum sample or a tongue swab sample, which would you choose? Please tell me why.

*Probe: What are the different reasons that your chose one sample over the other?*

Self-Collection vs. Health Worker Collected

17. If you had the choice of having the health worker swab your tongue or for you to swab your tongue yourself with the health worker supervising, which would you choose and why?

*Probe: Tell me about any concerns would you have about swabbing your own tongue?*

*Probe: Do you think it would be easy or hard to swab your own tongue? Why?*

18. If you were told you would need to swab your tongue yourself under the health workers supervision, how would you feel about it?

*Probe: What do you think about the possibility of taking the swab yourself without supervision?*

Trust and False-negative and False-positive Results

19. Please describe anything that makes you trust or not trust the use of tongue swabs for TB testing?
20. Would you be willing to have a tongue swab used for TB testing for yourself in the future? Why or why not.
21. There is no test that is always able to detect TB correctly every time. All tests have the possibility of saying a person does not have TB (a negative test result) when they do have TB. Also, some tests require samples that are more uncomfortable or harder to provide (meaning, it would be possible you could not provide a sample at all and TB could not be confirmed if you had TB). If you were offered the choice between a tongue swab test that has a slightly higher chance of missing TB when you have TB, and a second test that was more likely to detect TB but might be more uncomfortable or harder to provide a sample for, which would you choose and why?
22. Although rare, there is a small chance that the result of a tongue swab test could say someone has TB (a positive result) when they really do not. If you learned you were told you had TB and were started on TB treatment, when you actually did not have TB, how would this make you feel about the tongue swab test?

*Probe: Please tell me about the ways that receiving a positive test result for TB when you did not actually have TB would impact your life?*

23. Is there any additional information or support you would like to have before deciding about whether to have a tongue swab test for TB?

Wrap-up Question

24. Do you have any other thoughts you would like to share with me about tongue swabs or sputum for testing for TB?

**Appendix B.1: Images of Tongue Swab and Sputum Samples**

Tongue Swab

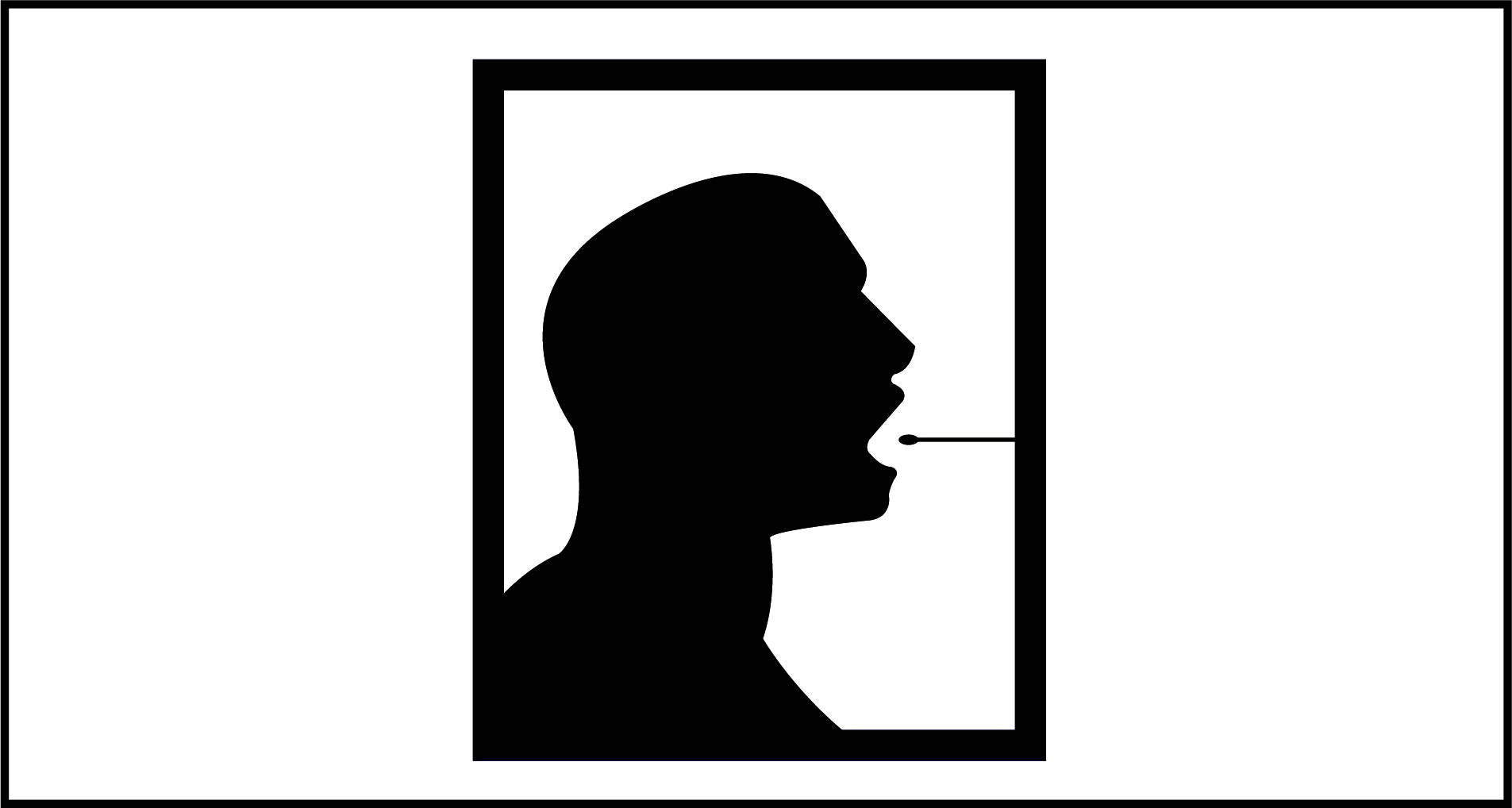

Sputum

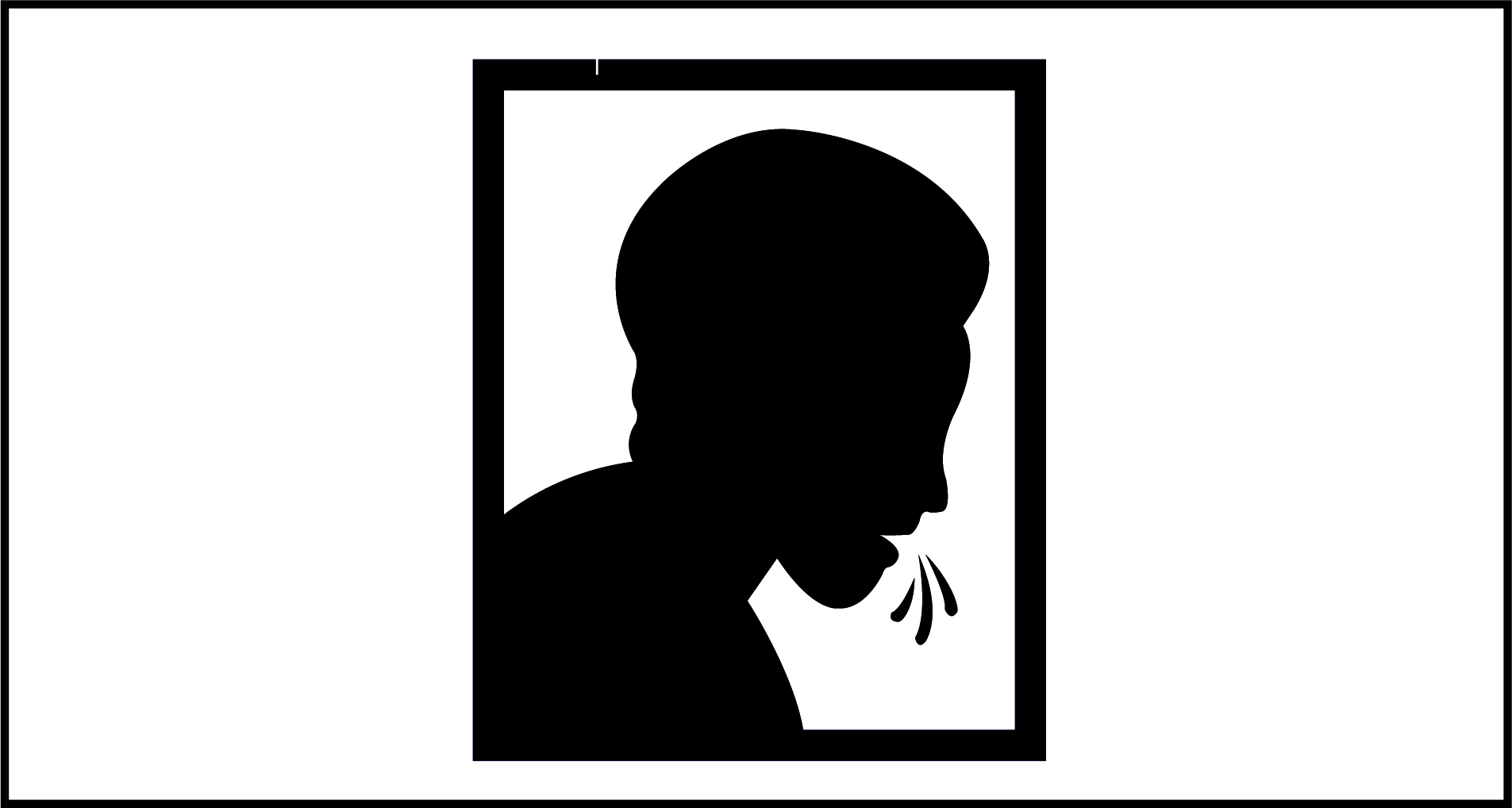

**Evaluating Acceptability and Usability for Tongue Swabs**

**Health Worker Questionnaire [to precede interview]**

| **To be completed by the interviewer** | |
| --- | --- |
| Date of interview | Free Form |
| Study Participant ID | Free Form |
| To ensure data integrity, please re-enter the Study participant ID. | Free Form |
| Name of country participant recruited from | 1. Zambia  2. Vietnam  3. Uganda  3. Philippines  4. India |
| What facility is the participant being recruited from (if more than one) | Select:   1. Local facility 1 2. Local facility 2 3. Local facility 3 |
| **Socio-demographics** |  |
| 1. What is your age? (in years) | Numeric (in years) |
| 2. What is your sex? | Select:   1. Male 2. Female |
| 3. What is your highest level of education started? | Select:   1. Never attended school 2. Primary school 3. Secondary school 4. Any tertiary (University or College) |
| 4. What type of health care worker are you? | Select:   1. Physician 2. Nurse 3. Pharmacist 4. Counsellor / treatment supporter 5. Other type of lay health worker |
| 5. How long have you worked in your current job/role? | Select:   1. Less than one year 2. More than one year but less than three years 3. More than three years but less than five years 4. Five years or more |
| 6. Have you ever been treated for TB disease in the past? | Select:   1. Yes 2. No |
| **Tongue Swab vs. Sputum Questions** |  |
| 7. Between sputum and tongue swab samples, which  is generally easier for you to obtain from a patient for TB diagnosis? | 1. Sputum  2. Tongue swab  3. No difference |
| 8. Would you prefer a health care provider to collect tongue swab samples from patients OR for the patients to collect the tongue swab sample under health care provider supervision? | 1. Health care provider  [Skip to Q10]  2. Patient under health care provider supervision  3. No preference [Skip to Q10] |
| 9. If you answered  that your preference is for the patient to collect the tongue swab sample under health care provider supervision how much does this relate to protecting healthcare providers from exposure to TB or other respiratory diseases? | 1. Very much  2. Some  3. A little  4. Not at all |
| 10. Overall, which sample do you prefer collecting from patients? | 1. Sputum  2. Tongue swab  3. No preference [End] |
| 11. [For those who answered sputum or tongue swab to the previous question]: Please tell me the why you prefer to collect this type of sample? [Select all that apply] | 1. Ease of collection  2. Comfort of the patient  3. Confidence in the result  4. The time it takes to collect or obtain the sample  5. Health worker or clinic staff safety |
| **Tongue Swab and Sputum Processing Questions** | |
| 12. On average, how much time do you spend per one patient for providing instructions about how to obtain a sputum sample? (This should include instructions before a patient is attempting to produce a sample and, if applicable, time spent with a patient while attempting to produce a sample. This should not include time spent with other patients or doing other tasks.) | 1. Less than 5 minutes 2. 5 minutes 3. 10 minutes 4. 15 minutes 5. 20 minutes 6. 30 minutes 7. Other: __________ 8. Can’t tell. |
| 13. On average, how long does it take for one patient, after having received prior instructions, to produce a sputum sample of acceptable quality? (This may include the time for possible repeated attempts to provide a sample in the case that the initial attempt was unsuccessful.) | 1. Less than 5 minutes 2. 5 minutes 3. 10 minutes 4. 15 minutes 5. 20 minutes 6. 30 minutes 7. Other: __________ 8. Can’t tell. |
| 14. On average, how much work time do you spend for processing one sputum sample received from a patient until it is ready for transport to the laboratory? (This should include the time for processing the sample but exclude administrative tasks such as filling in laboratory request forms. This may also include the work time for preparing samples for storage before transport but exclude the time a sample is stored/waiting until transport.) | 1. Less than 5 minutes 2. 5 minutes 3. 10 minutes 4. 15 minutes 5. 20 minutes 6. 30 minutes 7. Other: __________ 8. Can’t tell. |
| 15. On average, how much work time do you spend per workday for attending (all) patients with presumptive TB for the purpose of obtaining sputum samples and for processing samples received? (This is a total estimate over an average workday.) | 1. Less than 1 hour 2. 1 hour 3. 2 hours 4. 4 hours (half a workday) 5. 6 hours 6. 8 hours (full workday) 7. Other: __________ 8. Can’t tell |
| We will now focus on average work time for obtaining and processing a swab sample.  16. On average, how much time do you spend per one patient for providing instructions about obtaining a swab sample? (This should not include time spent with other patients or doing other tasks.) | 1. Less than 5 minutes 2. 5 minutes 3. 10 minutes 4. 15 minutes 5. 20 minutes 6. 30 minutes 7. Other: __________ 8. Can’t tell. |
| 17. On average, how long does it take to obtain a swab sample from a patient, after having provided prior instructions? | 1. <1 min 2. 1 minute 3. 2 minutes 4. 3 minutes 5. 5 minutes 6. Other: __________ 7. Can’t tell. |
| 18. On average, how much time do you spend for processing one swab sample obtained from a patient until it is ready for transport to the laboratory? (This should include the time for processing the sample itself to prepare for transport but exclude administrative tasks such as filling in laboratory request forms. This may also include the work time for preparing samples for storage before transport but exclude the time a sample is stored until transport.) | 1. Less than 5 minutes 2. 5 minutes 3. 10 minutes 4. 15 minutes 5. 20 minutes 6. 30 minutes 7. Other: __________ 8. Can’t tell. |
| 19. On average, how much work time per day do you think you would save if swab-based TB testing would completely replace sputum-based testing? | 1. Less than 1 hour 2. 1 hour 3. 2 hours 4. 3 hours 5. 4 hours (half a workday) 6. Other: __________ 7. Can’t tell. |
| 20. If so, what would be the most important ways by which swab-based TB testing saves time? [Select all that apply] | 1. No time savings 2. Less time required for instructions 3. Less time required for producing/obtaining a sample 4. Less time required for sample processing until it can be transported 5. Other: __________ 6. Can’t tell. |

**Topic guide for key informant interviews - health workers**

**Version 1.0 07 August 2023**

Background information (interviewer record information in fields below):

10. Facility recruited from:
11. Interview location:
12. Interview date (day/month/year):
13. Name of person conducting interview:

Introduction

Welcome to our interview today, we are grateful that you have volunteered to participate. We are conducting interviews with health workers like you who do TB-testing to learn about their experiences with or thoughts about two different types of samples that are collected and used to test people for TB. Overall, the goal is to understand the acceptability, usability, and preferences for a sample for TB testing that uses a tongue swab or sputum. There is no right or wrong answer. Please feel free to express your views during this interview. Your responses will be kept private and not shared with anyone else – only myself and the research team will read them. Your answers will not be linked to your name and will not affect your employment in any way. If there are any questions that you are uncomfortable answering, we can skip to the next question or stop the interview. Before we start, do you have any questions?

As a reminder, this interview will be audio recorded. I am going to start the recording now.

Warm-up Question

25. To start, please tell me about your job here at the clinic, including your role in testing people for TB.

*Probe: In your current job at this clinic do you do your work in the clinic, in the community, or both? Please tell me about the different places you work and what you do there.*

Experiences, Perceptions, and Acceptability

26. Currently at this clinic, people with symptoms of TB are given sputum to test for TB. We are also evaluating tongue swabs as part of a study on TB testing and so, as you know, tongue swabs for TB testing are being offered to patients as part of the study. Please tell me what you think about these two types of samples for TB testing: tongue swabs and sputum.

*Probe: Do you have any worries or concerns about either sputum or tongue swabs?*

27. Please tell me about your experience collecting tongue swab samples. Please tell me about this for both adults and children.

*Probe: Is it easy or hard to collect tongue swab samples? Why?*

*Probe: Tell me about what you like or dislike about the tongue swab*

*Probe: Do you experience any challenges collecting tongue swabs?*

*Probe: Do you find tongue swabs to be an acceptable way to collect a sample from a patient for TB testing?*

28. Please tell me about your experience collecting sputum samples. Please tell me about this for both adults and children.

*Probe: Is it easy or hard to collect sputum swab samples? Why?*

*Probe: Tell me about what you like or dislike about the sputum samples for TB testing?*

*Probe: Did you experience any challenges collecting sputum?*

*Probe: Do you find sputum to be an acceptable way to collect a sample from a patient for TB testing?*

16. Please tell me your overall thoughts on using tongue swabs for children compared to adults?

29. In your opinion, which type of sample (sputum or tongue swab) is easier for patients to provide? Please tell me about this for both adults and children.

*Probe: Which type of sample do you think patients prefer?*

30. In your opinion, which type of sample (sputum or tongue swab) would you rather have patients provide? Please tell me why. Please tell me about this for both adults and children.

Alternative phrasing: If you were counselling a patient and you could only recommend one type of TB sample collection to them (tongue swab or sputum), which would you choose and why?

*Probe: What are the reasons that your chose one sample over the other?*

31. Please tell me about concerns, if any, that you have with collecting a sputum sample for TB testing?

*Probe: Does personal safety/exposure to TB impact your feelings about a sputum sample?*

32. Please tell me about concerns, if any, with collecting a tongue swab sample for TB testing?

*Probe: Does personal safety/exposure to TB impact your feelings about a tongue swab sample?*

33. If both sputum and tongue swab were available in the healthcare setting where you work and recommended for TB testing, which would you prefer as a first test for individuals with TB symptoms/presumptive TB? Please tell me why.

*Probe: Are there times you would be more likely to use tongue swabs? What about sputum?*

*Probe: Do you feel differently about this for adults as compared to children?*

Collection

34. If you had the choice of collecting a tongue swab from the patient vs. the patient swabbing their own tongue to collect the sample, which would you choose and why?

*Probe: Tell me about any concerns would you have about swabbing their tongue?*

35. Please tell me how you think, if at all, using tongue swabs as opposed to collecting sputum would impact your day-to-day work here at the clinic?

*Probe: In what ways would using tongue swabs as opposed to sputum change the amount of time you spend supporting TB testing throughout the day?*

*Probe: Please tell me about any aspects of tongue swabs for TB testing that could cause difficulties in its day-to-day use in your setting.*

Trust and False-negative and False-positive Results

36. Please describe anything that makes you trust or not trust the use of tongue swabs for TB testing?

*Probe: Do you have any concerns about using a tongue swab test? Please tell me about them.*

37. If you needed to be tested for TB, would you want to have a tongue swab or provide sputum? Please tell me why.
38. If your child were being tested for TB in the future, would you want them to have a tongue swab or provide sputum? Please tell me why.
39. Thinking about both tongue swabs and sputum, which do you feel more confident in having a patient provide? Please tell me about this for both adults and children.
40. Imagine two types of tests for TB: one is easy to collect, including from children, but has a slightly lower chance of actually detecting TB in someone who has TB. The other test is better at detecting TB in someone who has TB, but is harder to collect, and not all people like children will be able to provide a sample using this more accurate test. Which would you choose and why?

*Probe: How do your feelings about these two tests change or differ when thinking about a patient who is an adult or adolescent, compared to a child.*

41. Although rare, there is a small chance that the result of a tongue swab test could say a patient has TB (a positive result) when they really do not. If this rare situation happened to a patient of yours and they were told they had TB when they actually did not, how would this make you feel?

*Probe: Please tell me about the ways that receiving a positive test result for TB when someone does not actually have TB would impact your patients?*

*Probe: Have you ever had this happen with a sputum test result? Please tell me about it.*

18. Is there any additional information or support you want to have before counselling your patients about tongue swab or sputum tests for TB?

Probe: What else would you want to know about tongue swabs before using them with your patients?

Wrap-up Question

19. Do you have any other thoughts you would like to share with me about tongue swabs or sputum for testing for TB?

**Appendix B.2: Images of Tongue Swab and Sputum Samples**

Tongue Swab

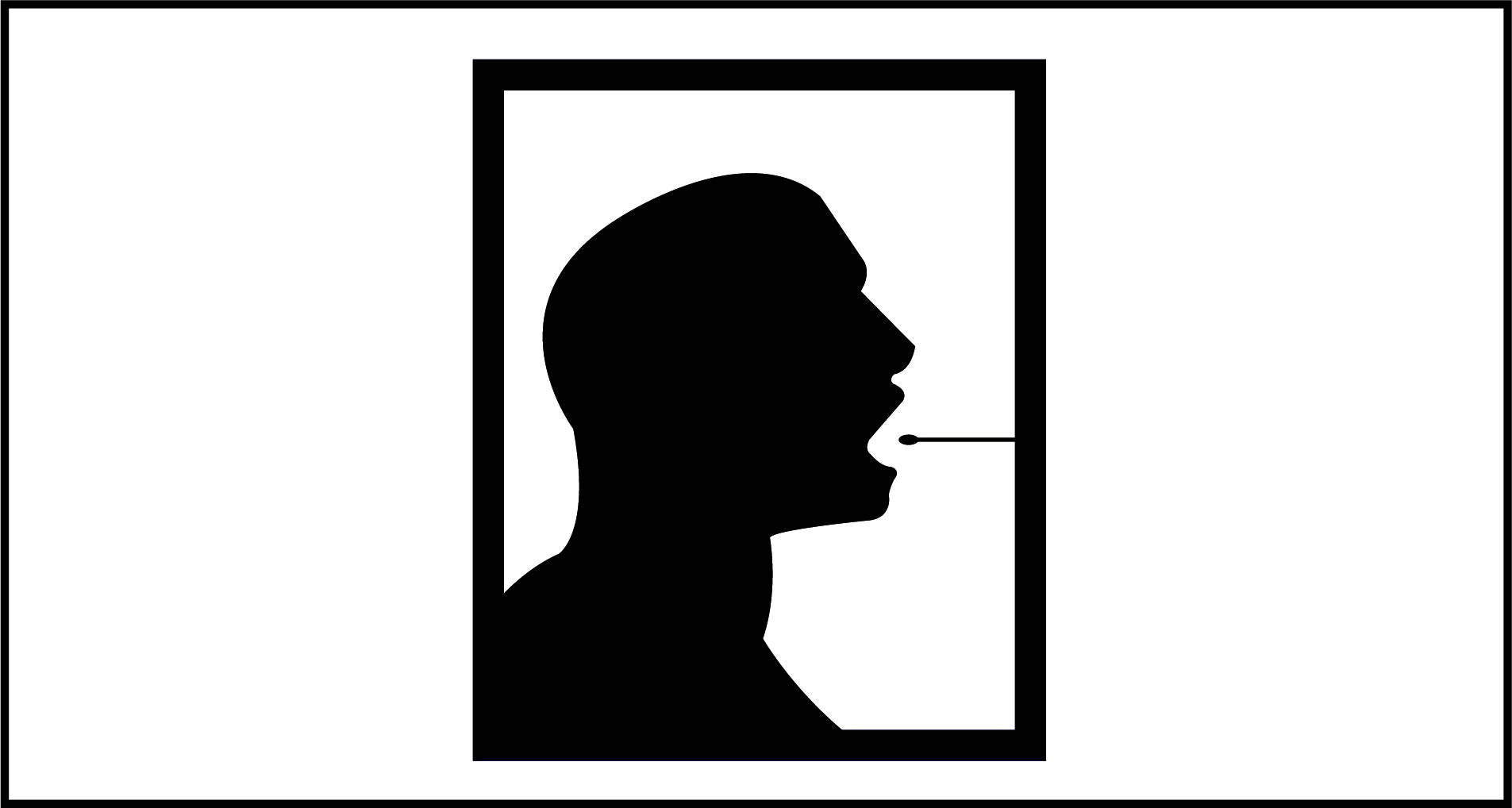

Sputum

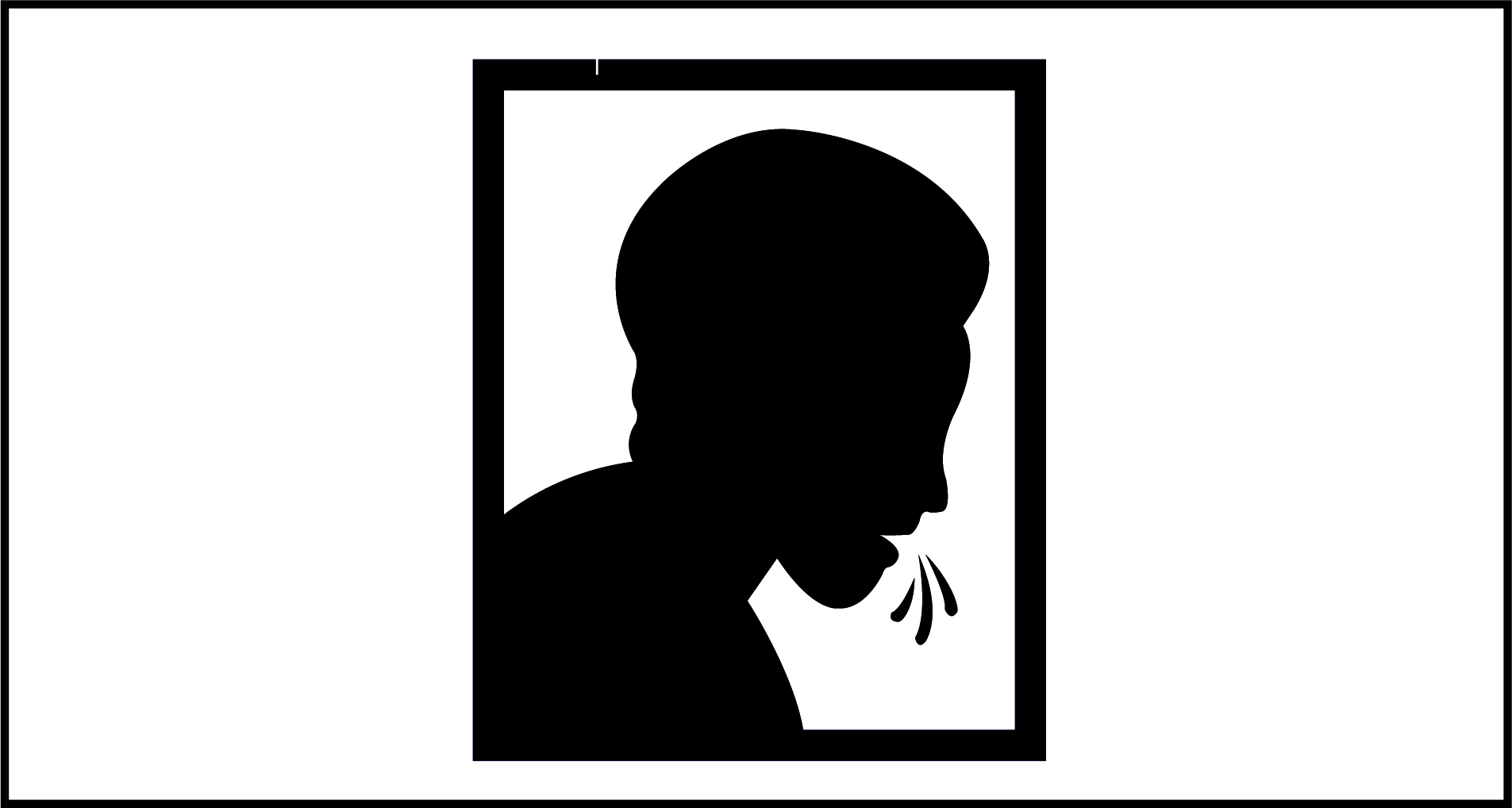

**Evaluating Acceptability and Usability for Tongue Swabs**

**Topic guide for semi-structured interviews: caregivers of children testing for TB**

**Version 1.0 07 August 2023**

Background information (interviewer record information in fields below):

14. Participant ID:
15. Participant type:
16. Facility recruited from:
17. Participant age (in years):
18. Participant sex:
19. Participant caregiver relationship to child being tested for TB:

Parent

Grandparent

Aunt/Uncle

Sibling

Other, Specify: ____________

20. Interview location:
21. Interview date (day/month/year):
22. Name of person conducting interview:
23. Consent obtained?	Yes	No (do not proceed with interview until informed consent is obtained)

Introduction

Welcome to our interview today, we are grateful that you have volunteered to participate. We are conducting interviews with people or their caregivers who came to the clinic and were offered testing for TB to learn about their experiences with or thoughts about two different types of samples that are collected and used to test people for TB. Overall, the goal is to understand the acceptability, usability, and preferences for a sample for TB testing using a tongue swab or sputum which is coughing a substance from the lungs into a cup. There is no right or wrong answer. Please feel free to express your views during this interview. Your responses will be kept private and not shared with any of the staff at your local clinic – only myself and the research team will read them. Your answers will not be linked to your name and will not affect the type of treatment or care you receive. If there are any questions that you are uncomfortable answering, we can skip to the next question or stop the interview. Before we start, do you have any questions?

As a reminder, this interview will be audio recorded. I am going to start the recording now.

Warm-up Question

42. To start, please tell me about the reasons you decided to come to the clinic today.

Experiences, Perceptions, and Acceptability

43. Today your child or the child you brought to the clinic was offered two types of testing for TB – sputum and tongue swabs. Please tell me what you think about these two types of tests.

*Probe: Do you have any worries or concerns about either test?*

44. Please tell me your thoughts about your child being tested for TB with a tongue swab. When I say tongue swab samples, I mean the swabs like this (show image – Appendix 1).

*Probe: Tell me about anything you like or dislike about tongue swabs.*

*Probe: Did you have any concerns about your child or the person you brought to the clinic today having a tongue swab test? Please tell me about it.*

**[For caregivers of participants who provided tongue swab samples]**

45. Please tell me about your child or the child you brought to the clinic’s experience with the tongue swab samples?

*Probe: Was it easy or hard for them to give the tongue swab sample? Why?*

*Probe: Tell me about what you or they liked or disliked about the tongue swab?*

46. Please describe, based on what they say, how your child felt physically when the health worker used the tongue swab?

*Probe: Did they experience any pain or discomfort?*

47. When the health worker swabbed your child’s tongue, please tell me what you thought about the instructions or information the health worker gave them before, during, or after collecting the sample?

*Probe: How did the health worker explain the tongue swab to you?*

*Probe: What concerns or questions did you have before, during, or after the procedure?*

*Probe: What information would you have liked to have about the tongue swab before, during, or after the procedure?*

**[For caregivers of participants who provided sputum samples]**

48. Please tell me about your child or the child you brought to the clinic’s experience with the sputum samples?

*Probe: Was it easy or hard for them to give the sputum sample? Why?*

*Probe: Tell me about what you or they liked or disliked about the sputum?*

49. Please describe, based on what they say, how your child felt physically when providing the sputum sample.

*Probe: Did they express or experience any pain or discomfort?*

50. When the health worker collected or explained the collection of the sputum sample, please tell me what you thought about the instructions or information the health worker gave them before, during, or after collecting the sample?

*Probe: How did the health worker explain the sputum sample to you?*

*Probe: What concerns or questions did you have before, during, or after the procedure?*

*Probe: What information would you have liked to have about the sputum sample before, during, or after the procedure?*

51. Please tell me about any concerns, if any, that you have with your child providing a sputum sample for TB testing?

**[For caregivers of children who did not provide tongue swab samples]**

52. Please tell me why you decided not to have your child give a tongue swab sample today?

*Probe: What did you think when the health worker asked to give you a tongue swab?*

*Probe: Please tell me about any worries or concerns you had about having your tongue swabbed today.*

**[For all caregivers]**

53. Please tell me about how you would feel if you brought your child to the clinic for TB testing and you were told they would have only a tongue swab to test for TB because it would be too hard to collect a sputum sample?

*Probe: Would having a tongue swab as the only test for your child make you feel uncomfortable or comfortable. Please tell me why.*

*Probe: Would you rather have a different test? Please tell me why.*

54. Please tell me about anything that would influence your choice to accept or decline/not accept tongue swabs for TB testing in the future for your child?
55. Please tell me about any problems your child experienced during or after the tests for TB were done that came from taking the test.

*Probe: Please tell me about anything that was difficult or inconvenient about the test they had.*

56. If you had a choice between your child providing a sputum sample or a tongue swab sample, which would you choose? Please tell me why.

*Probe: What are the different reasons that your chose one sample over the other?*

Self-Collection vs. Health Worker Collected

57. If you had the choice of having the health worker swab your child’s tongue or for you to swab their tongue yourself with the health worker supervising, which would you choose and why?

*Probe: Tell me about any concerns would you have about swabbing their tongue?*

*Probe: Do you think it would be easy or hard to swab their tongue? Why?*

58. If you came to the clinic and were told to swab your child’s tongue under the health workers supervision, how would you feel about it?

Trust and False-negative and False-positive Results

59. Please describe anything that makes you trust or not trust the use of tongue swabs for TB testing?
60. Would you be willing to have a tongue swab used for TB testing for yourself in the future? Why or why not.
61. There is no test that is always able to detect TB correctly every time. All tests have the possibility of saying a person does not have TB (a negative test result) when they do have TB. Also, some tests require samples that are more uncomfortable or harder to provide (meaning, it would be possible your child could not provide a sample at all and TB could not be confirmed if your child had TB). If you were offered the choice for your child between a tongue swab test that a slightly higher chance of missing TB when they had TB, and a second test that was more likely to detect TB but might be more uncomfortable or harder to provide a sample for, which would you choose and why?
62. Although rare, there is a small chance that the result of a tongue swab test could say a child has TB (a positive result) when they really do not. If you learned your child was told they had TB and was started on TB treatment, when they actually did not have TB treatment, how would this make you feel about the tongue swab test?

*Probe: Please tell me about the ways that receiving a positive test result for TB when you did not actually have TB would impact your life?*

22. Is there any additional information or support you would like to have before deciding about your child having a tongue swab test for TB?

Wrap-up Question

23. Do you have any other thoughts you would like to share with me about tongue swabs or sputum for testing for TB?

**Appendix B.3: Images of Tongue Swab and Sputum Samples**

Tongue Swab

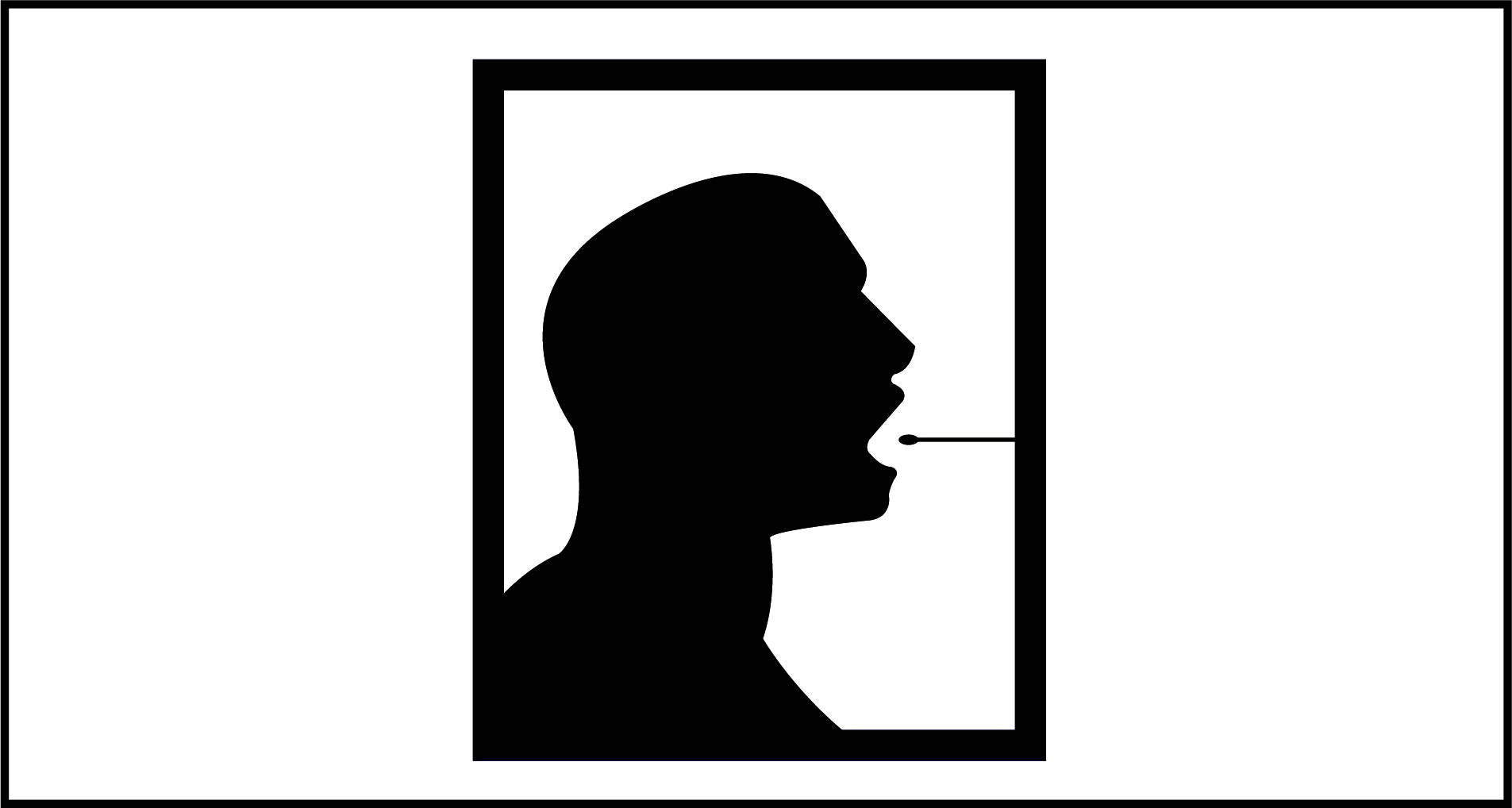

Sputum

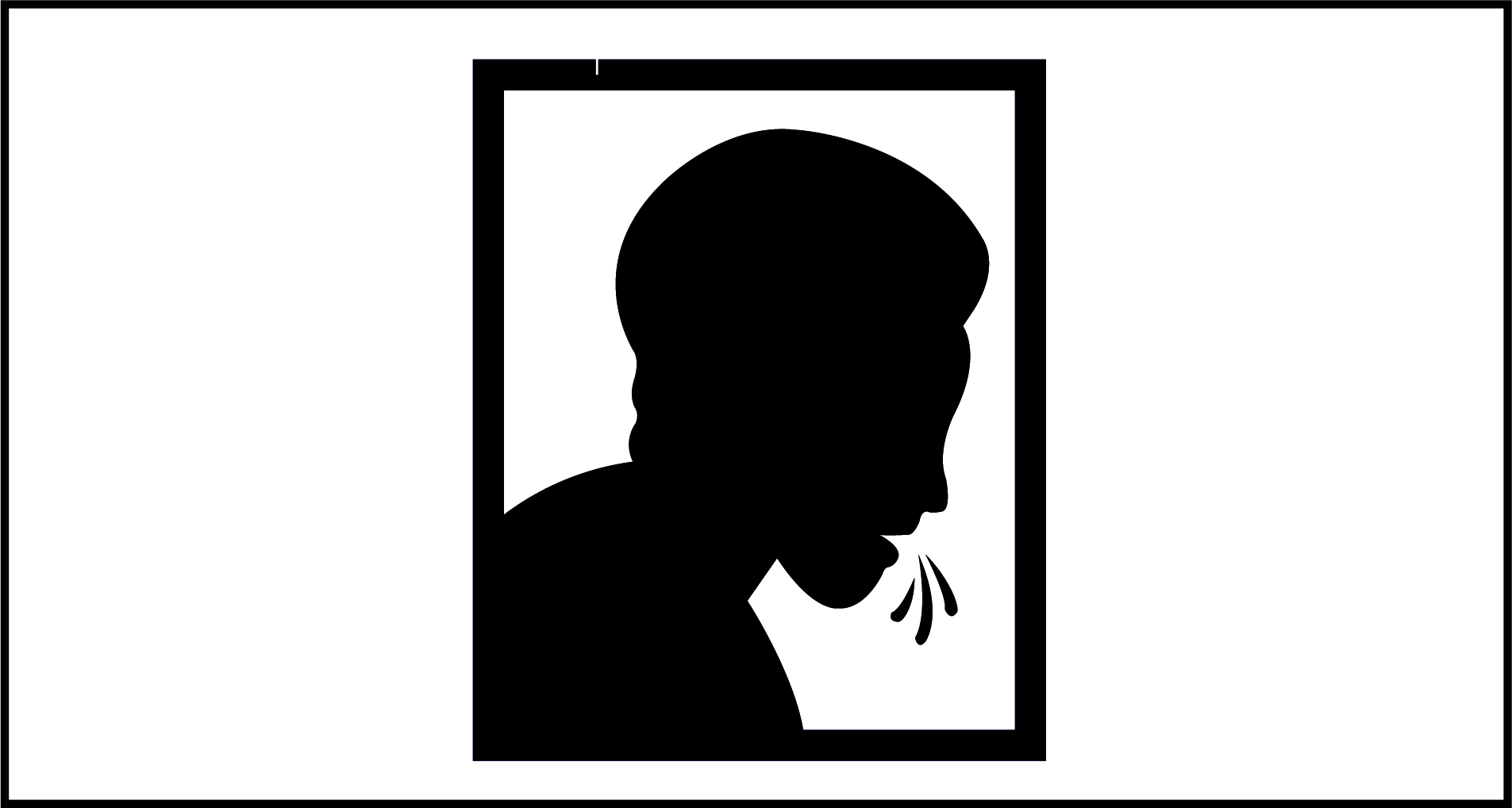
